# Maternal prenatal psychological distress is associated with aortic intima-media thickness in infants in the FinnBrain Birth Cohort Study

**DOI:** 10.1101/2024.06.27.24309083

**Authors:** Anna-Katariina Aatsinki, Martha Snäll, Katja Pahkala, Hasse Karlsson, Laura Perasto, Olli T. Raitakari, Linnea Karlsson, Suvi P. Rovio

## Abstract

**Aims:** The origins of cardiovascular health are embedded in early life. It has been suggested that exposure to prenatal stress is linked with markers of cardiovascular structure and functioning. We aimed to explore in the FinnBrain Birth Cohort Study if and how maternal prenatal psychological distress – a potentially modifiable risk factor – associates with infant aortic intima-media thickness (aIMT).

**Methods:** Maternal symptoms were measured 2-3 times during pregnancy using EPDS for depressive symptoms, SCL-90 anxiety subscale for general anxiety and PRAQ-R2 for pregnancy-specific anxiety. Vascular ultrasonography was performed at the age of 2-3 months (n=273) and aIMT was measured.

**Results:** In adjusted models, mean PRAQ-R2 across pregnancy was positively associated with aIMT. In boys, mean EPDS across pregnancy was associated with a thicker aIMT.

**Conclusions:** Our results indicate that pregnancy-specific anxiety associates with thicker aIMT in both sexes, whereas maternal depressive symptoms associate with thicker aIMT in boys only.

**Lay Summary:** Stress during pregnancy changes maternal physiology, and is related with child development. We studied how mother’s tress, i.e. symptoms of depression and anxiety, during pregnancy links with cardiovascular structure of the infants.

- Mothers were asked about their symptoms of depression and anxiety 2-3 times during pregnancy. They were also asked about pregnancy-specific anxiety, meaning worries related to labour, future health of the child and pregnancy-related changes. At the age of 2-3 months, infants were imaged with ultrasound, and the thickness of two innermost layers of the wall of abdominal aorta was measured (aIMT). The thickness may relate with cardiovascular disease risk.
- pregnancy-related anxiety was linked with thicker aIMT. Additionally, boys showed thicker aIMT when their mothers had depressive symptoms during pregnancy.

## Introduction

Atherosclerotic cardiovascular diseases are major contributors to disability and mortality worldwide^1^. The origins of atherosclerosis are embedded in early life and exposure to atherosclerosis risk factors in childhood associates with increased risk for cardiovascular diseases in later life^2^. In addition to the well-established risk factors such as elevated blood pressure, adverse serum lipid profile and smoking, increasing evidence shows that psychosocial stress is implicated in the development of various forms of atherosclerotic cardiovascular diseases^3^. Additionally to stress in adulthood, childhood adverse psychosocial factors are associated with cardiovascular diseases and related risk factors in adulthood^4^ . Furthermore, prior evidence suggests that early life stress may associate with the risk of atherosclerotic cardiovascular diseases even when accounting for the traditional risk factors^5^. It has been shown that childhood adverse psychosocial factors are associated with carotid intima-media thickness^5^, which is a marker of early atherosclerosis in adults^2^. In children, intima-media thickness of the abdominal aorta may be a more sensitive marker of subclinical atherosclerosis than carotid intima-media thickness because structural changes in the aorta occur before those in the carotid arteries^6^. A recent systematic review and meta-analysis examined the association between exposures during the first 1000 days of life and aortic intima-media thickness measured from fetuses and children up to 10 years. The study identified that having been born small or large for gestational age, intrauterine growth restriction, preeclampsia, and maternal smoking all associated with greater aortic intima-media thickness in neonates^7^.

Although stress in childhood and adulthood is associated with atherosclerotic cardiovascular diseases, less is known of prenatal stress^4,5^. Prenatal stress encompasses different exposures such as prenatal stressful events, maternal prenatal psychosocial distress as well as physiological markers of stress^8^. Importantly, prenatal stress is known to be associated with greater morbidity^9,10^. Prenatal stress can lead to altered hypothalamus pituitary adrenal axis functioning^11^, affect functioning of the placenta^12^, increase inflammation^13^ and cause epigenetic alterations^14,15^, all of which may participate in the prenatal programming of health and disease^8^. Furthermore, although intima-media thickness has not been studied in prenatally stressed animals, animal studies have indicated that prenatal stress exposure increases the atherosclerotic plaque size especially in aged mice^16,17^, but not in younger conspecifics^17,18^. In humans, studies focusing on the associations between exposure to prenatal stressful events, such as wartime bombings and famine, and atherosclerotic cardiovascular diseases have yielded mixed results^19,20^, most likely owing to the diverse nature of the exposures and outcomes.

This study was set out to investigate the links between prenatal stress exposure and very early markers of cardiovascular disease risk, i.e. infant aortic intima-media thickness. With very early assessment of aortic intima-media thickness, we aimed to characterize the influence of prenatal rather than postnatal factors. Moreover, we aimed to study maternal prenatal psychological distress, which is a prevalent form of prenatal stress in western populations^21,22^. More specifically, we wanted to study the maternal depressive as well as general and pregnancy-specific anxiety symptoms across pregnancy and their associations with infant aortic intima-media thickness. We hypothesized that maternal psychological distress during pregnancy associates with thicker aortic intima-media in the infant. Additionally, it is well established that prenatal stress exposure may have sex-specific effects^23^ on various outcomes such as asthma^24,25^ and coronary heart disease risk score^26^, and therefore, we also explored possible effect modification by child’s biological sex assigned at birth.

## Methods

### Study Sample

This study is a part of the ongoing longitudinal FinnBrain Birth Cohort Study focusing on the combined influence of environmental and genetic factors on child development and later health outcomes^27^. For this study, a subsample of the FinnBrain Birth Cohort Study mothers, fathers and their infants were recruited based on the age of the infant (i.e. 6-8 weeks at the time of the study visit) and available information on prenatal distress. The participants were contacted via telephone and offered a possibility to take part in the ultrasonography subcohort. The data collection took place between January 2014 and January 2016. Overall, 327 infants attended the study visit. The subsample included two twin pairs, although only on one of these pairs ultrasonography was successfully completed on both siblings. Ultrasonography was successfully acquired from 273 infants. The maternal prenatal psychological distress scores were similar regardless of ultrasonography success (Wilcoxon rank-sum test, Edinburgh Postnatal Depression Scale p-value = 0.22, Symptom Checklist-90, anxiety subscale p-value = 0.365, Pregnancy Anxiety Questionnaire-Revised p-value = 0.332).

The study has been approved by the Ethics Committee of the Hospital District of Southwest Finland. Written informed consent was obtained from all participating mothers and fathers, and mothers provided written informed consent on behalf of their infants.

### Prenatal psychological distress

Prenatal maternal psychological distress symptoms were queried on gestational weeks 14, 24, and 34 using the Symptom Checklist-90, anxiety subscale (SCL)^28,29^, pregnancy anxiety questionnaire-revised (PRAQ-R2)^30^, and the Edinburgh Postnatal Depression Scale (EPDS)^31,32^. EPDS and SCL were used to assess general depressive and anxiety symptoms, respectively, whereas PRAQ-R2 was used to measure pregnancy-related worries and anxiety related to the fear of giving birth, worries about bearing a physically or mentally handicapped child, and concerns about the mother’s own appearance. PRAQ-R2 was included in the first measurement point (gestational weeks 14) only later in the data collection sweep, and this resulted in a lower sample size available for the PRAQ-R2 in the first measurement point (Tables 1 and 2).

**Table 1.**
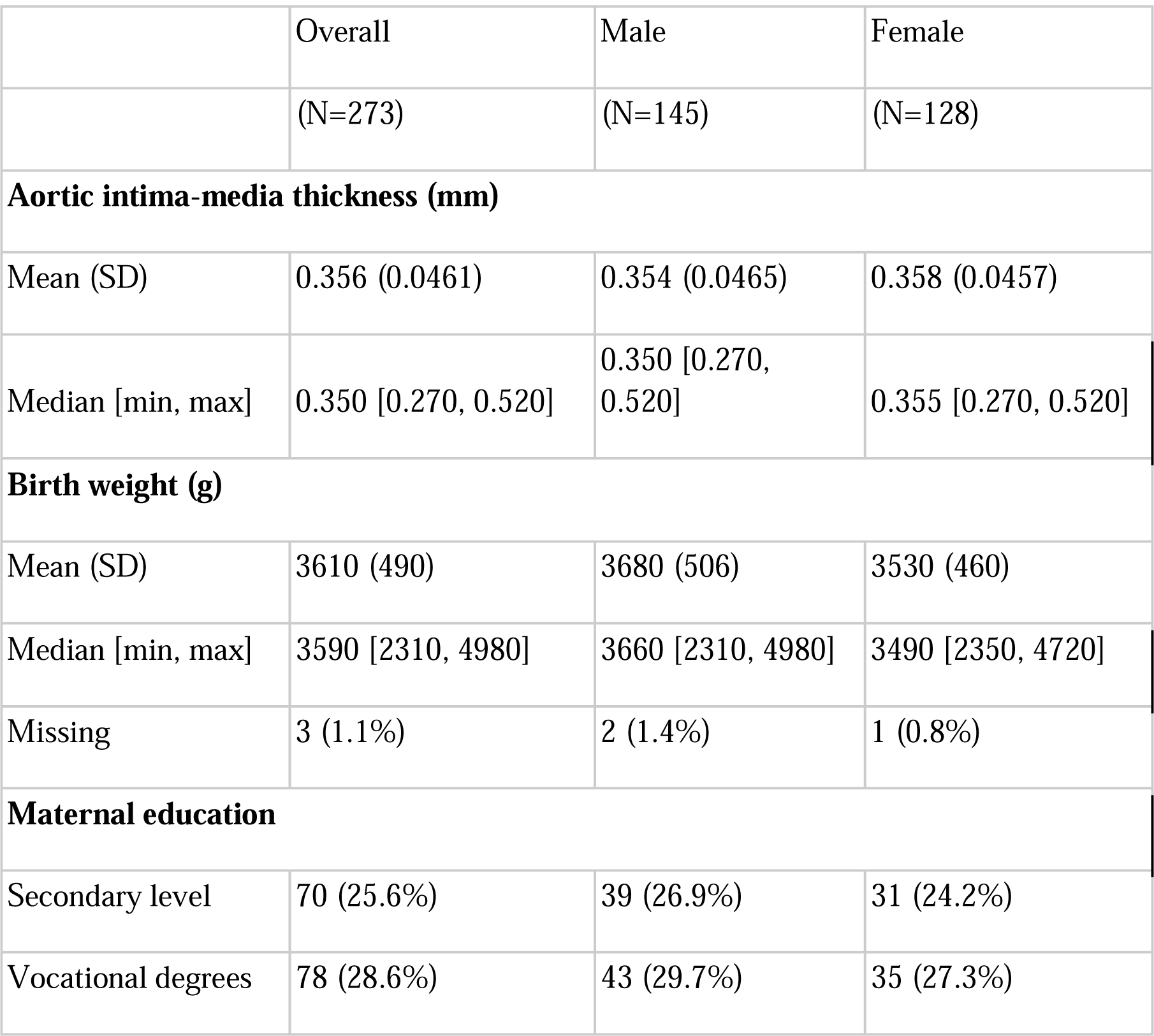
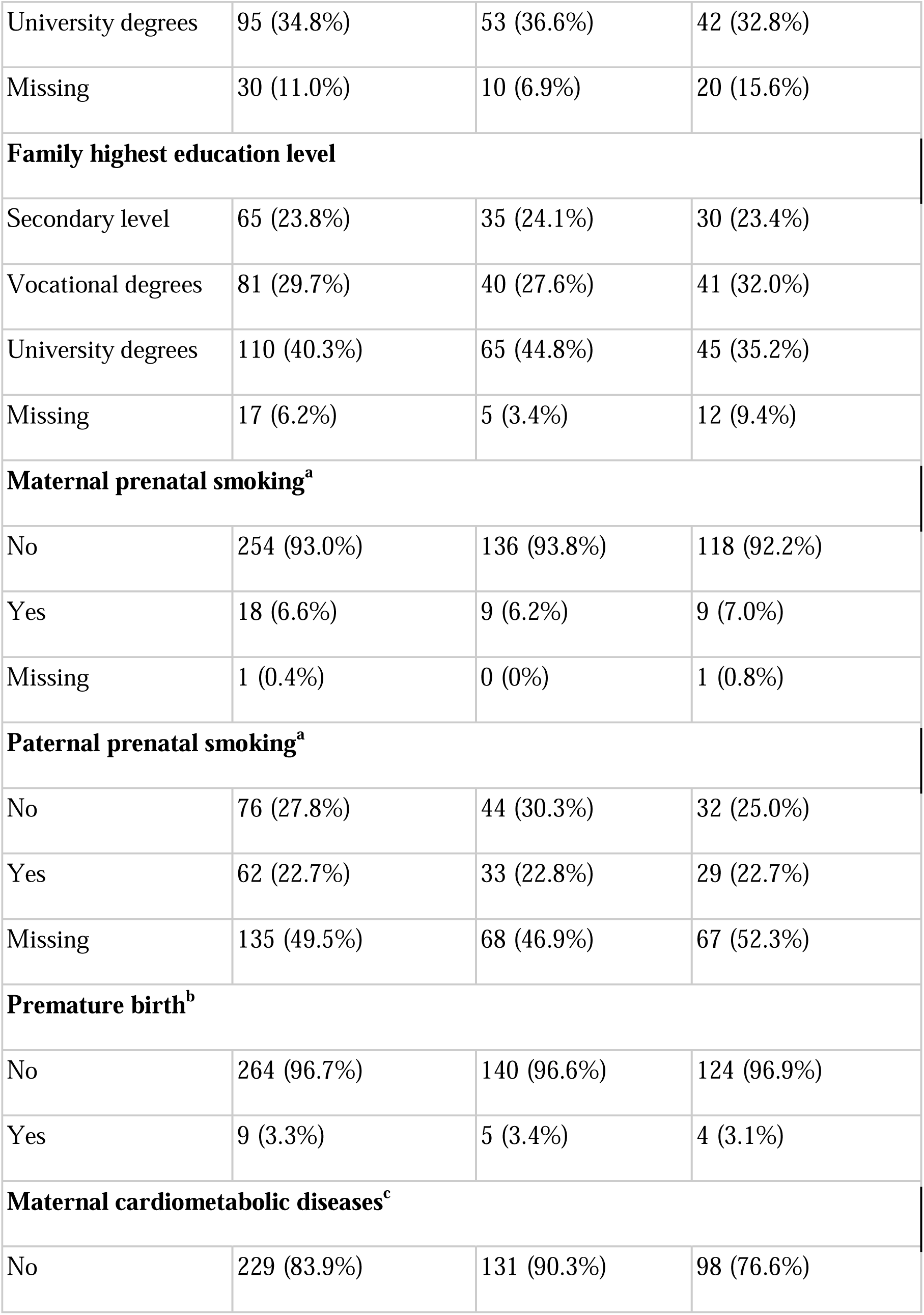
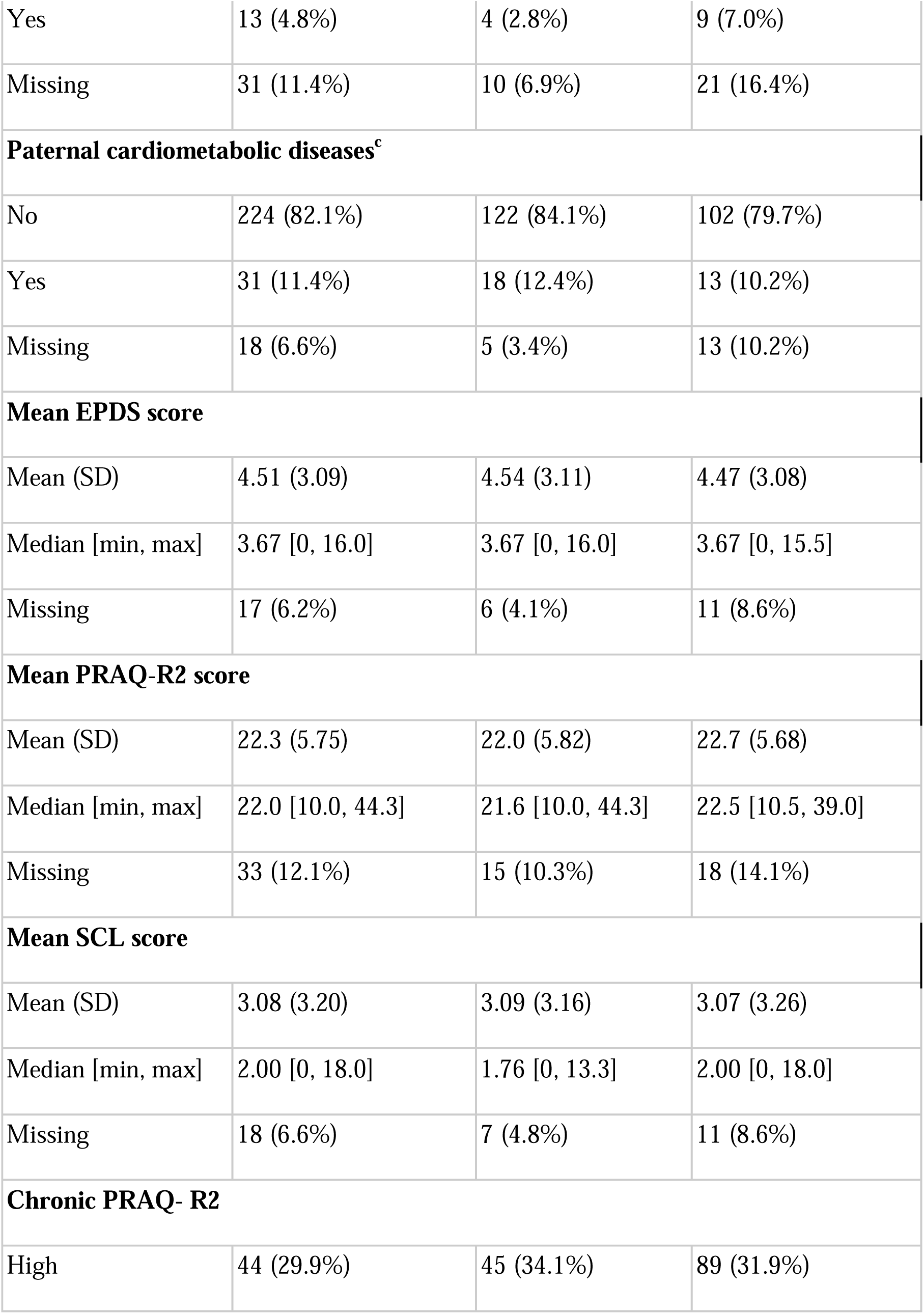
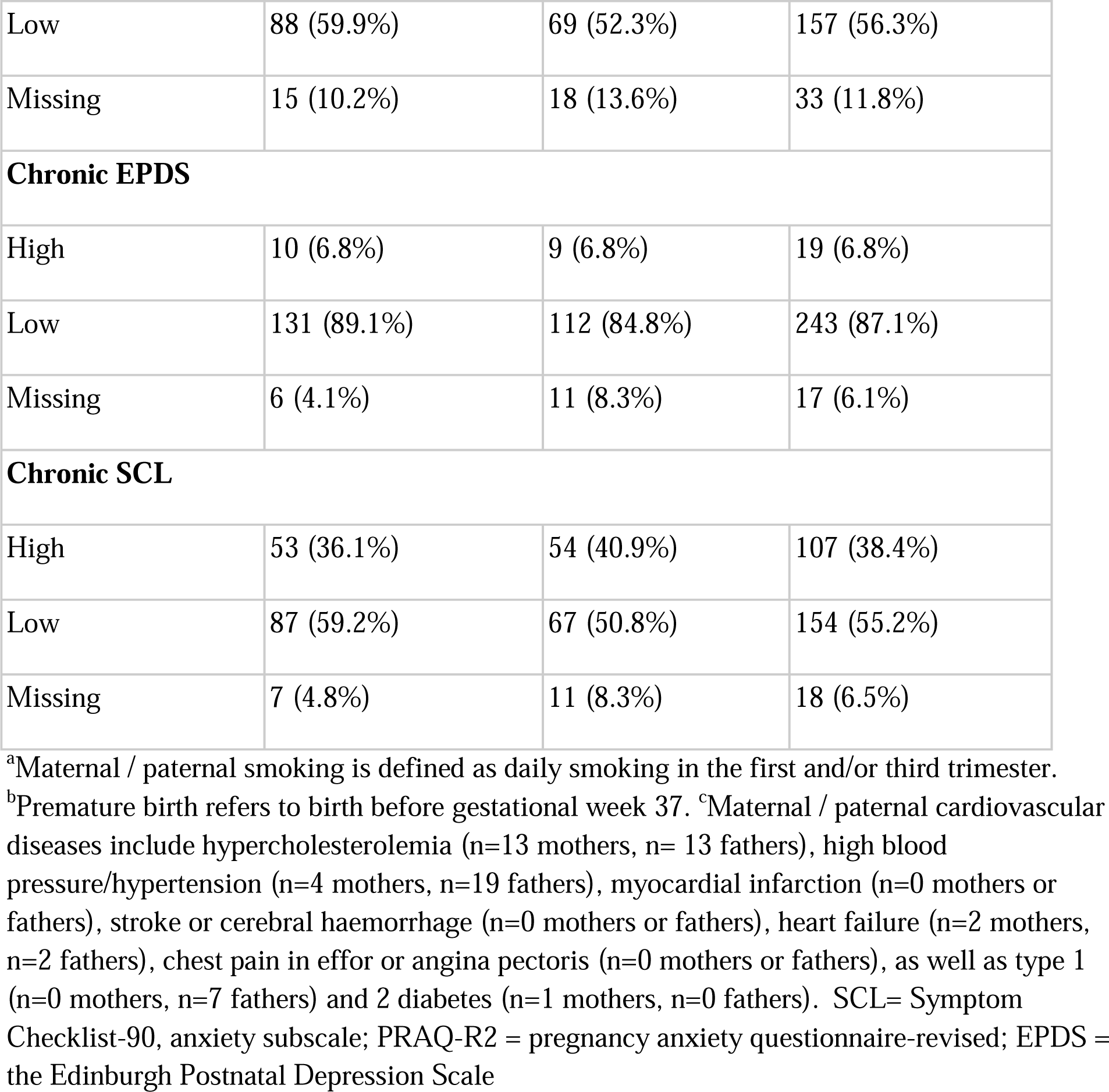
The overall sample characteristics.

**Table 2.**
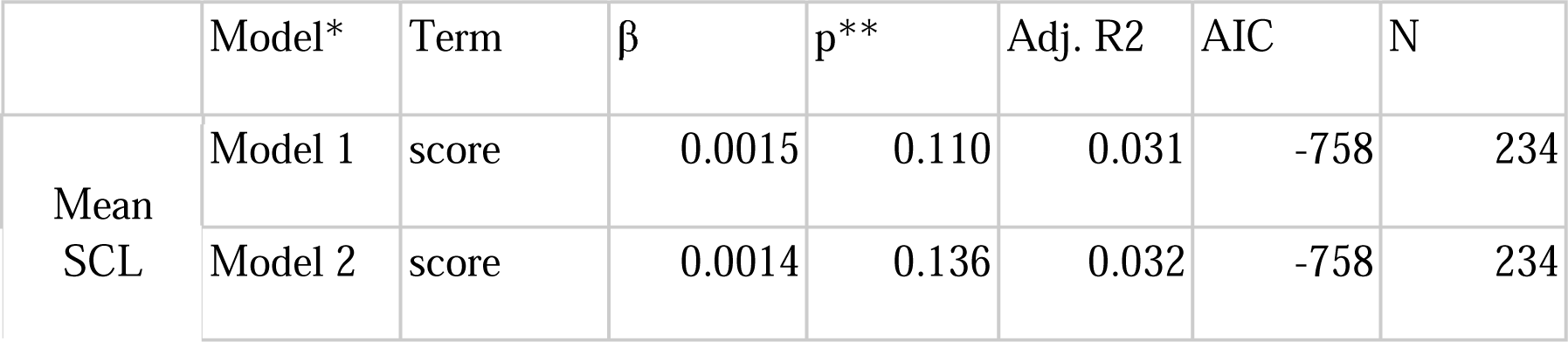
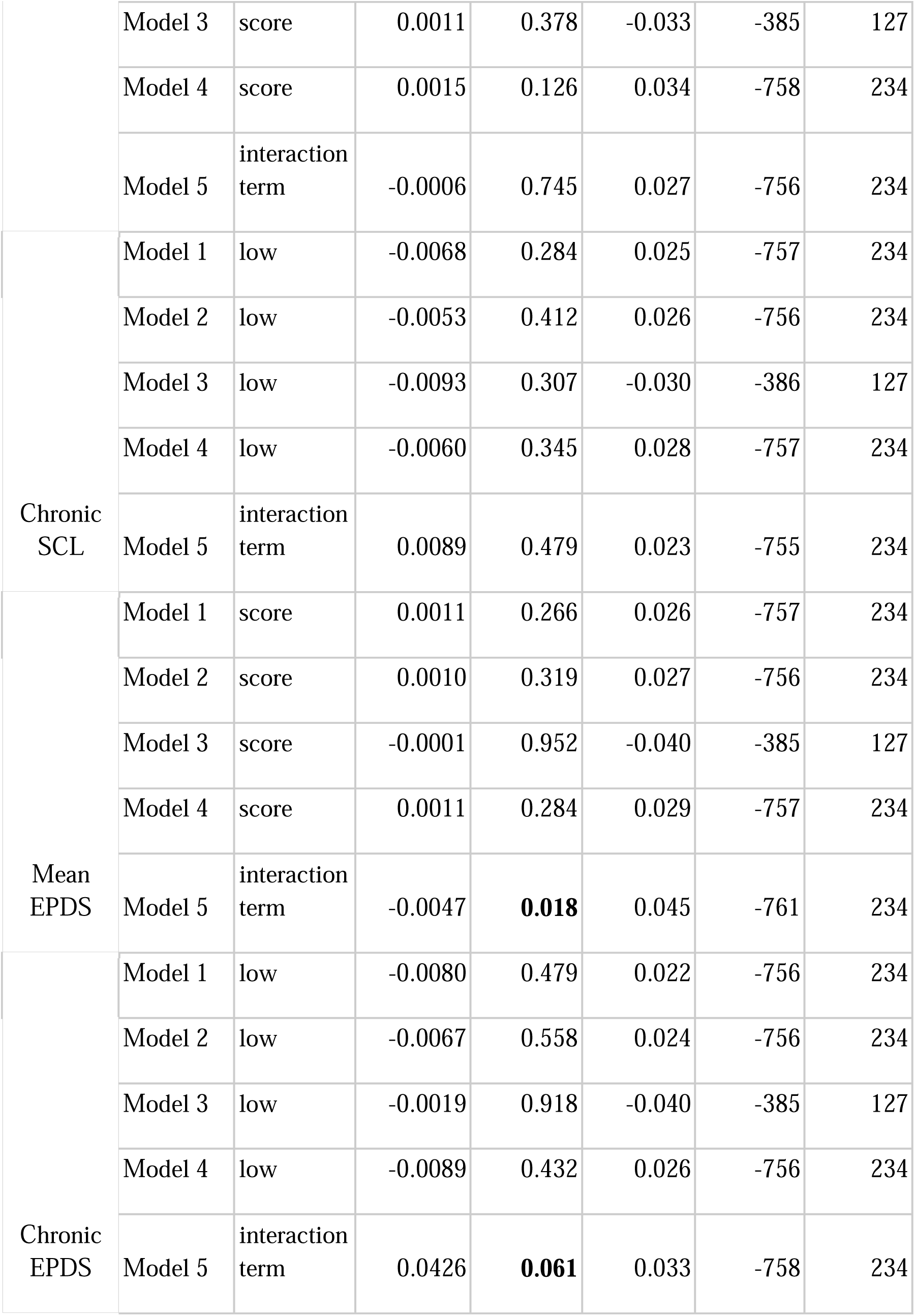
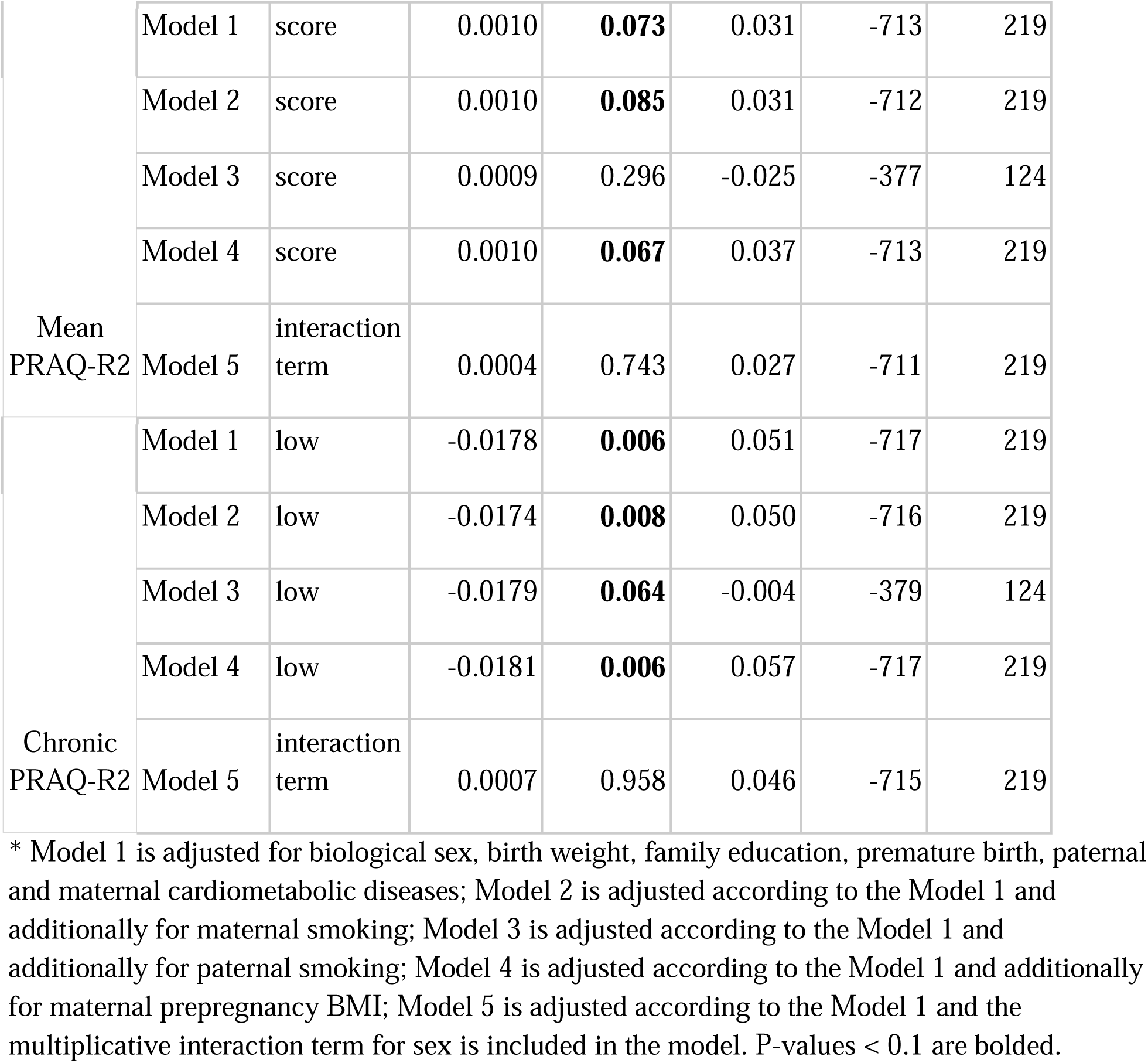
Results from step-wise regression models. The mean variables are continuous values, whereas constantly high variables are binary.

| | Model* | Term | $\beta$ | p** | Adj. R2 | AIC | N |
| --- | --- | --- | --- | --- | --- | --- | --- |
| Mean<br>SCL | Model 1 | score | 0.0015 | 0.110 | 0.031 | -758 | 234 |
|  | Model 2 | score | 0.0014 | 0.136 | 0.032 | -758 | 234 |
|  | Model 3 | score | 0.0011 | 0.378 | -0.033 | -385 | 127 |
|  | Model 4 | score | 0.0015 | 0.126 | 0.034 | -758 | 234 |
|  | Model 5 | interaction term | -0.0006 | 0.745 | 0.027 | -756 | 234 |
| Chronic SCL | Model 1 | low | -0.0068 | 0.284 | 0.025 | -757 | 234 |
|  | Model 2 | low | -0.0053 | 0.412 | 0.026 | -756 | 234 |
|  | Model 3 | low | -0.0093 | 0.307 | -0.030 | -386 | 127 |
|  | Model 4 | low | -0.0060 | 0.345 | 0.028 | -757 | 234 |
|  | Model 5 | interaction term | 0.0089 | 0.479 | 0.023 | -755 | 234 |
| Mean EPDS | Model 1 | score | 0.0011 | 0.266 | 0.026 | -757 | 234 |
|  | Model 2 | score | 0.0010 | 0.319 | 0.027 | -756 | 234 |
|  | Model 3 | score | -0.0001 | 0.952 | -0.040 | -385 | 127 |
|  | Model 4 | score | 0.0011 | 0.284 | 0.029 | -757 | 234 |
|  | Model 5 | interaction term | -0.0047 | **0.018** | 0.045 | -761 | 234 |
| Chronic EPDS | Model 1 | low | -0.0080 | 0.479 | 0.022 | -756 | 234 |
|  | Model 2 | low | -0.0067 | 0.558 | 0.024 | -756 | 234 |
|  | Model 3 | low | -0.0019 | 0.918 | -0.040 | -385 | 127 |
|  | Model 4 | low | -0.0089 | 0.432 | 0.026 | -756 | 234 |
|  | Model 5 | interaction term | 0.0426 | **0.061** | 0.033 | -758 | 234 |
| Mean<br>PRAQ-R2 | Model 1 | score | 0.0010 | **0.073** | 0.031 | -713 | 219 |
|  | Model 2 | score | 0.0010 | **0.085** | 0.031 | -712 | 219 |
|  | Model 3 | score | 0.0009 | 0.296 | -0.025 | -377 | 124 |
|  | Model 4 | score | 0.0010 | **0.067** | 0.037 | -713 | 219 |
|  | Model 5 | interaction<br>term | 0.0004 | 0.743 | 0.027 | -711 | 219 |
| Chronic<br>PRAQ-R2 | Model 1 | low | -0.0178 | **0.006** | 0.051 | -717 | 219 |
|  | Model 2 | low | -0.0174 | **0.008** | 0.050 | -716 | 219 |
|  | Model 3 | low | -0.0179 | **0.064** | -0.004 | -379 | 124 |
|  | Model 4 | low | -0.0181 | **0.006** | 0.057 | -717 | 219 |
|  | Model 5 | interaction<br>term | 0.0007 | 0.958 | 0.046 | -715 | 219 |

\* Model 1 is adjusted for biological sex, birth weight, family education, premature birth, paternal and maternal cardiometabolic diseases; Model 2 is adjusted according to the Model 1 and additionally for maternal smoking; Model 3 is adjusted according to the Model 1 and additionally for paternal smoking; Model 4 is adjusted according to the Model 1 and additionally for maternal prepregnancy BMI; Model 5 is adjusted according to the Model 1 and the multiplicative interaction term for sex is included in the model. P-values < 0.1 are bolded.

All symptom scales resulted in continuous variables. As we wanted to study thepotentially accumulating xposure over the course of pregnancy, we created new variables summarizing the symptoms over pregnancy. Based on the continuous variables, we created two separate outcome variables for each questionnaire: 1) a mean variable over each available prenatal measure, henceforth mean EPDS, SCL and PRAQ-R2, and 2) a categorical variable using a cut-off point per time point and indicating constantly high exposure, henceforth constantly high EPDS, PRAQ-R2, or SCL. The total score of 10 or more was used as a cut-off point for EPDS to present more clinically significant, although mild to moderate symptoms^32^. Median split was used for the other questionnaires since we are not aware of clearly defined cut-off points. For the constantly high exposure variables, mothers who scored above a cut-off point in two measurement points were classified as having constantly high scores in the given questionnaire. Subsequently, mothers with less than two measures were excluded from the analyses. In this sample, the mean SCL and EPDS correlated moderately (Spearman ρ=0.7), whereas PRAQ correlation with EPDS and SCL was smaller (ρ=0.35 for EPDS, ρ=0.33 for SCL).

### Ultrasonography

The infants underwent ultrasonography examination of abdominal aorta at a study visit at age 6-8 weeks. Ultrasonography was performed in a silent and dark room using Siemens Acuson Sequoia (Acuson, Mountain View, CA, USA) ^33^. For the aortic intima-media thickness measurements, the most distal part of the abdominal aorta wall was scanned, and the image was focused on the far wall (i.e. dorsal arterial wall). Images that were 15 mm in width were magnified using a resolution box function. Several clips were taken during heart cycles for blinded offline analyzes. Using ultrasonic calipers, at least 4 measurements covering the entire far wall segment of interest during end-systole were taken for each image and the average of these measurements was used as the mean aortic intima-media thickness (mm). Offline image processing was performed using ComPACS 10.7.8 (MediMatic Solutions, Genova, Italy) analysis program.

### Covariates

The child’s sex, birth weight and prematurity as well as family socioeconomic status (defined as highest education level of the parents), parental cardiometabolic diseases, and exposure to mother’s and father’s tobacco smoking during pregnancy were used as potential confounders of the studied associations (Fig 1.). Child’s biological sex was assigned at birth based on outer genitalia inspection, hereafter referred as sex. Information on the child’s birth weight (g) and duration of gestation (preterm < 37 gestational weeks) were collected from the National Birth Registry provided by the National Institute for Health and Welfare (www.thl.fi). Information on parents’ education, own and partners cardiometabolic diseases, and parents’ smoking were collected using questionnaires on gestational weeks 14, 24 and 34. Parent’s education was categorized into 1) secondary (high school or vocational education), 2) vocational university (university of applied sciences) and 3) university (bachelor, master or doctoral degree). Both parents’ education level was considered, and the higher education level was used as a measure for the family’s socioeconomic status. Cardiometabolic conditions included self-reported hypercholesterolemia (n=13 mothers, n=13 fathers), high blood pressure/hypertension (n=4 mothers, n=19 fathers), myocardial infarction (n=0 mothers or fathers), stroke or cerebral haemorrhage (n=0 mothers or fathers), heart failure (n=2 mothers, n=2 fathers), chest pain in effort or angina pectoris (n=0 mothers or fathers), as well as type 1 (n=0 mothers, n=7 fathers) and 2 diabetes (n=1 mothers, n=0 fathers) measured with binary questions. Additionally, both parents were categorized into smokers and non-smokers according to their self-reported information on current daily smoking. If fathers had not enrolled in the study or if they had not reported their smoking habits, we used the mothers’ reports on their partner’s smoking habits.

**Figure 1.**
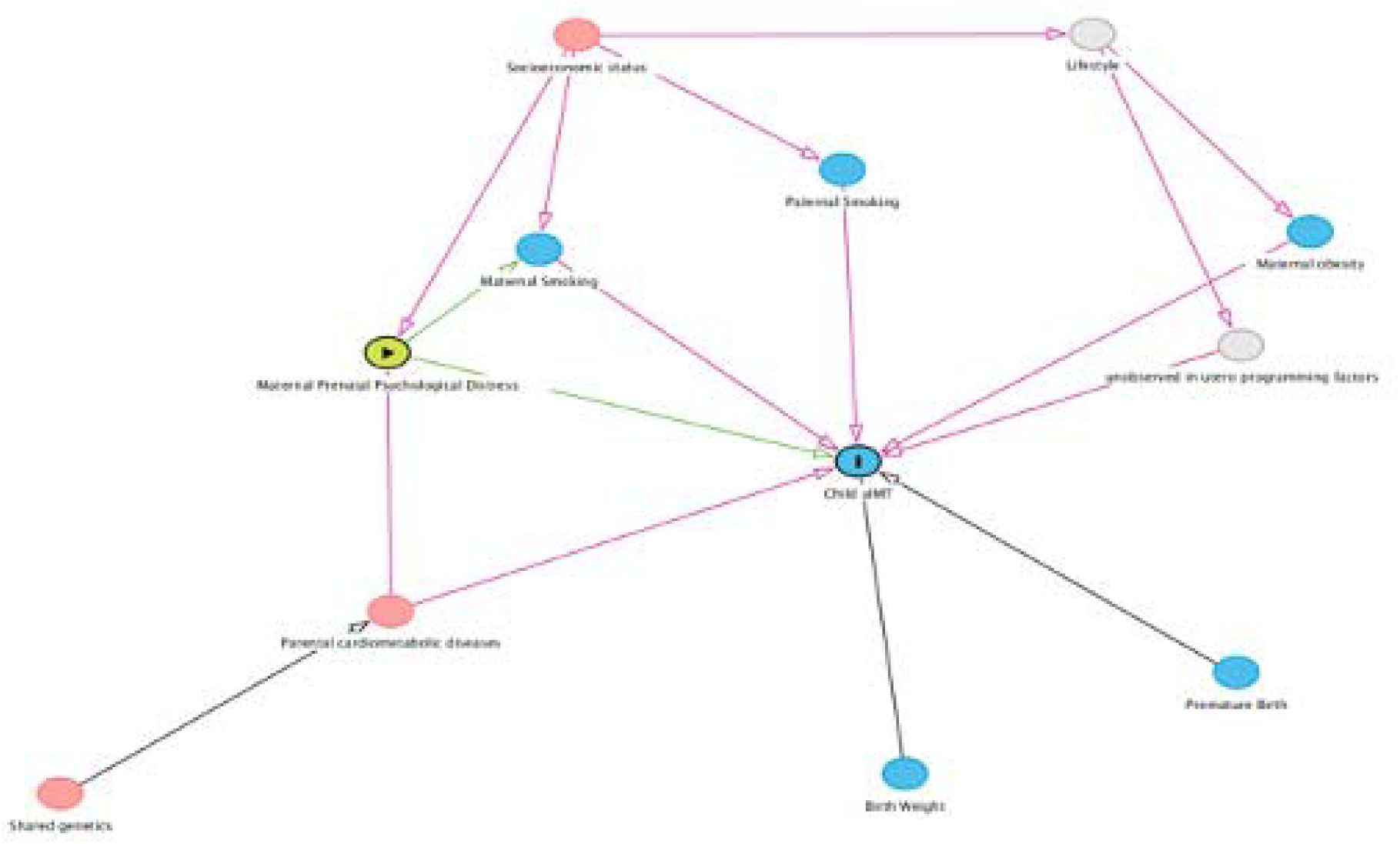
Directed Acyclic Graph on the causal model. DAGitty identified parental cardiometabolic diseases and socioeconomic status as minimal sufficient adjustment sets for estimating the total effect of maternal prenatal psychological distress on child aortic intima-media thickness (aIMT). exposure; outcome; ancestor of exposure; ancestor of outcome; ancestor of exposure *and* outcome; unobserved (latent); causal path; biasing path. Black lines indicate a path that was not identified as causal or biasing paths.

Nearly all mothers provided data on prenatal smoking while these data were missing from half of the fathers (Table 1). SCL and EPDS had higher sample size (n=234) than the analyses regarding PRAQ-R2 (n=218, Table 2). Based on the availability of covariates and psychological symptom reports, sample sizes in the final adjusted analyses varied between 124 and 234 (Table 2).

### Statistical analyses

Infant aortic intima-media thickness was the dependent variable and maternal constantly high and mean EPDS, SCL and PRAQ-R2 were treated as independent variables in the statistical analyses. We performed linear regression models with stepwise adjustments. Stepwise adjustment was done due to varying number of missing observations in the covariates. Directed Acyclic Graph with DAGitty was used to illustrate the causal model and to detect the relevant covariates (Fig 1). The linear models were adjusted for child’s sex, birth weight, gestational weeks, family SES, and cardiometabolic conditions reported by the parents (model 1). Subsequently, we added to the model tentative mediators such as maternal or paternal smoking (model 2 for maternal and model 3 for paternal smoking), maternal prepregnancy obesity (model 4) to the model 1 one at a time. As prenatal psychological distress may associate with child outcomes in sex-specific manner, we also adjusted all analyses for child’s sex at birth, and finally, introduced a ‘sex*psychological distress’ interaction term into the model without mediators (i.e. model 1) resulting in a separate statistical model (model 5). Models were built separately for the mean and constantly high psychological distress score variables. The analyses were conducted in R (R Core team, version 4.3.2).

## Results

The general characteristics of the sample with ultrasonography resemble the overall FinnBrain Birth Cohort Study sample and reflect the overall population of Southwestern Finland (Table 1)^27^. In a stepwise adjusted model, constantly high PRAQ-R2 was directly associated with infant aortic intima-media thickness (p-value 0.055-0.008, Table 2, Fig2*a,* 2*b*). In a similar direction, mean PRAQ-R2 scores were directly associated with aortic intima-media thickness, but the associations did not reach conventional statistical significance (p-values 0.067-0.300, Table 2). When adjusting for paternal smoking with a high number of missing values (49.5 %, Table 1), the p-value attenuated for both variables, but the direction and magnitude of the point estimates remained the same (constantly high PRAQ-R2 p=0.064, mean PRAQ-R2 p=0.300, Table 2). SCL or EPDS did not have an association with infant aortic intima-media thickness (Fig2*c*, 2d, 2*e*, 2*f*). However, we observed that both mean EPDS and constantly high EPDS showed sex-interaction (p-value 0.02 and 0.06, Table 2, Figure 2*d*). In sex-specific analyses, boys had a direct association between mean EPDS scores and aortic intima-media thickness (β=0.003, p=0.012) while no association was observed among girls. Moreover, aortic intima-media was thicker in boys of mother’s scoring constantly high in EPDS, although this association did not reach the conventional statistical significance (p=0.073).

**Figure 2.**
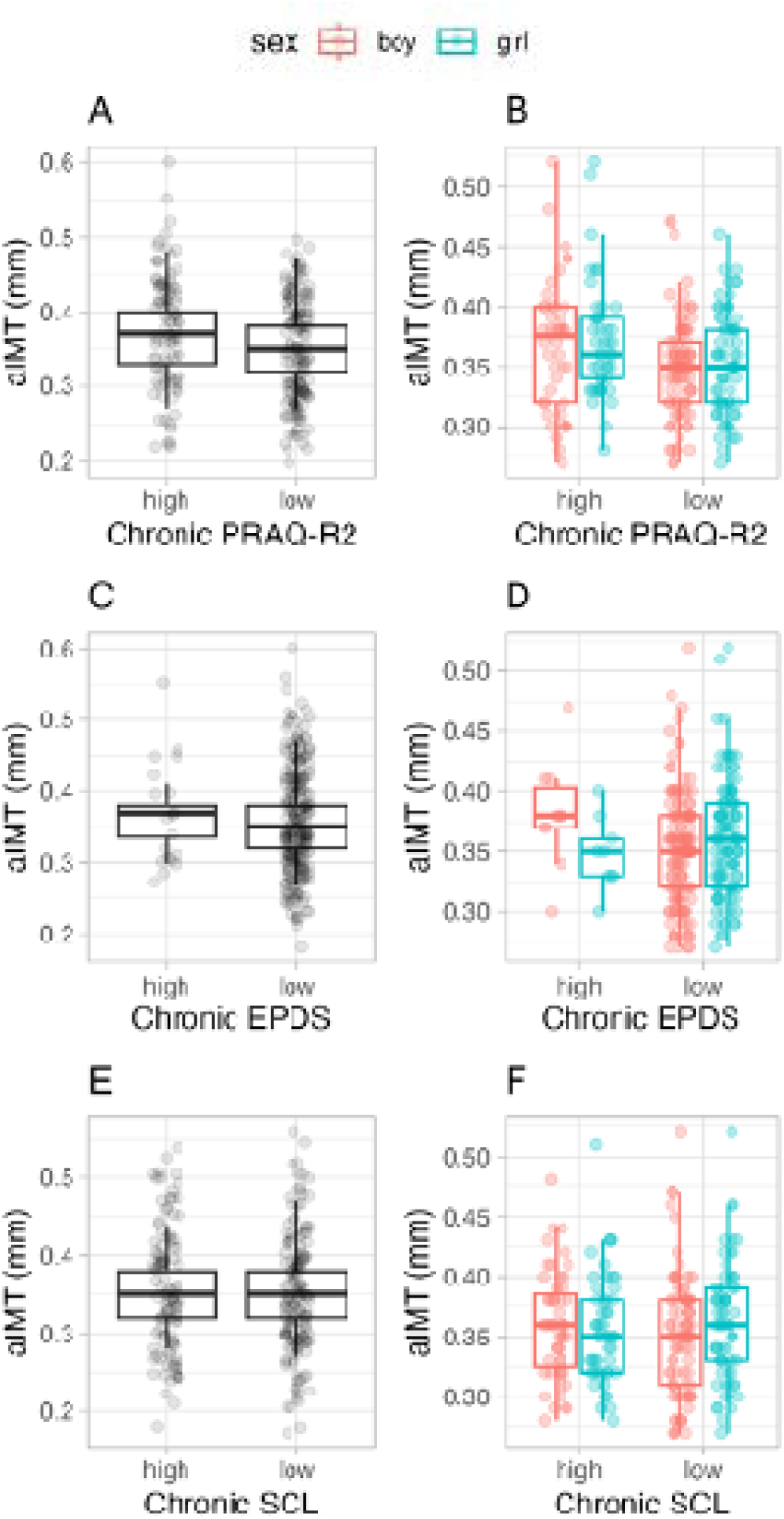
Constantly high EPDS in A) the whole population and B) by sex. Constantly high SCL in C) the whole population and D) by sex. Constantly high PRAQ-R2 in E) the whole population and F) by sex. Chronic = mother scores over the cut-off point in two measurement points; no = mother did not score above the cut-off point. aIMT = aortic intima-media thickness

As a sensitivity analysis, we tested the association between maternal symptoms and aortic intima-media thickness in a subsample of data with all covariate and symptom data available (n=124, Supplementary Table 1). Regarding constantly high PRAQ-R2, the estimate direction and magnitude remained similar, although the association was not statistically significant (p-values >0.063 for all associations, Supplementary Table 1). Regarding mean PRAQ-R2 scores, the direction of the estimate remained similar, but again the p-values and magnitude of estimates attenuated (p-values >0.28 for all associations, Supplementary Table 1).

## Discussion

It is suggested that exposure to different forms of stress both prenatally, in childhood and through life increases the risk for cardiovascular diseases^4,5,20,26^. To our knowledge, our study is the first to report associations between mild to moderate maternal prenatal psychological distress and infant aortic intima-media thickness, a marker of subclinical atherosclerosis in adults^34^. We showed that both pregnancy-specific anxiety and depressive symptoms were associated with thicker infant aortic intima-media. However, the association with prenatal depressive symptoms was sex-specific, and symptoms of depression were associated with thicker aortic intima-media only in boys. The very early assessment of aortic intima-media thickness indicates largely an influence of prenatal stress or alternatively a common cause, such as shared heritable background of the phenotypes. Previous papers have identified having been born small or large for gestational age, intrauterine growth restriction, and preeclampsia as risk factors for thicker neonatal intima-media, but other determinants of neonatal and infant intima-media are less studied. Our work adds on the very early determinants of infant intima-media thickness.

Previous studies on prenatal stress exposure and cardiovascular diseases have yielded mixed results, which is partially due to variance in the definitions and measures of stress. Plana-Ripoll and colleagues reported in a Danish register-based study that prenatal stress, i.e. bereavement, may be associated with increased risk of heart disease and hypertension at the age of 40^20^. The association attenuated when the shared postnatal environment of the siblings was considered, indicating that the finding might be confounded by postnatal factors. On the other hand, Schreirer et al. have reported from a Finnish register-based sample that prenatal exposure to bombings during wartime was associated with higher coronary heart disease survival rate in females suggesting a protective role of stress exposure during unstable societal time^19^. Along with the disparities in the published results, one must acknowledge that maternal prenatal psychological distress is a distinct stress exposure from stressful life events, and different exposures may have differential impact on pregnancy physiology and child outcomes^35^.

Cortisol is the end-product of the hypothalamus pituitary adrenal -axis and the main regulator of stress response^8^. Besides stressful life events, maternal high cortisol concentrations have been associated with child’s outcomes such as thicker carotid intima-media at the age of 1 year^36^, higher blood pressure at the age of 3 years^37^ as well as lower arterial elasticity at the age of 5-7 years^38^. These findings indicate that hypothalamus pituitary adrenal -axis activity is potentially participating in development of atherosclerosis, reflected in intima-media thickness. On the other hand, a previous study suggested that exposure to maternal prenatal psychological distress was not associated with blood pressure in childhood nor the vascular function and structure measured as arterial distensibility, stiffness, and brachial diameter^39,40^. The previous results potentially indicate that it would be hypothalamus pituitary adrenal -axis alteration per se, but not necessarily mild to moderate levels of maternal psychological distress as such, which is associated with blood pressure and vascular function and structure during childhood.

A body of literature has indicated that exposure to prenatal stress excerpts sex-specific effects on pregnancy and a range of child outcomes such as asthma, neuropsychological and psychiatric symptoms^23^. However, previous studies on cardiovascular outcomes have often overlooked the sex-specificity of the findings, while these outcomes present with sex differences as well. Here, we showed that maternal depressive symptoms, but not general or pregnancy-related anxiety symptoms, associated with intima-media thickness in a sex-specific manner. Infant boys exposed to maternal constant mild to moderate depressive symptoms were more likely to have thicker intima-media in comparison with the non-exposed babies, which would indicate that males are more susceptible to vascular effects after prenatal maternal depressive symptoms than females. Depressive symptoms during pregnancy may associate differently with physiological changes in the mother compared with pregnancy-specific and general anxiety^11^. We have demonstrated this in a partially overlapping sample from the FinnBrain Birth Cohort Study by showing that chronic and increasing maternal depressive symptoms are associated with elevated long-term hair cortisol concentrations in pregnant women while no associations with pregnancy-specific or general anxiety and cortisol concentrations were observed^11^. However, Stinson et al. have linked prenatal cortisol exposure with higher cardiovascular risk score in adulthood only in females^26^. Otherwise, we are not aware of prior reports on sex-specific findings related to maternal prenatal psychological distress and infant cardiovascular outcomes. Although, our observations and those of previous studies indicate sex-specific influences of prenatal stress exposure, comparability of the findings is somewhat hampered by differences in exposure and outcome definitions and timings.

In addition to the dysregulation of the hypothalamus pituitary adrenal -axis, maternal prenatal psychological distress may exert effects on offspring vascular development via other mechanisms, such as immune system aberration, epigenetic alterations and changes in placental physiology^41–44^. A recent study suggested that pregnancy-specific anxiety may be linked with increased pro-inflammatory cytokine balance^45^. Moreover, fetal abdominal aorta wall has been found to contain macrophages and other inflammatory elements in the context of intrauterine growth restriction, which is coupled with intima-media thickening^46^. Similarly, amniotic fluid of fetuses with growth retardation and intima-media thickening has elevated proinflammatory cytokine concentrations^47^. However, future mechanistic and biomarker studies are needed to elucidate the likely mechanism behind our observation.

Although we only addressed maternal psychological distress, and not e.g. stressful life events, as a stress exposure, our study benefited from multiple measurement points through pregnancy. This enabled us to study more chronic patterns of stress that may have physiological impact^11^, especially when the study sample is drawn from general population in which the prevalence of psychosocial stressors is relatively low as compared with cohorts drawn from e.g. populations with socioeconomic difficulties, substance use or psychiatric illnesses. Moreover, the aortic intima-media thickness was assessed as early as in age 6-8 weeks, which may diminish the potential effects of postnatal confounders. However, long-term follow-up would be beneficial to determine the longitudinal associations and relevance for later health. Moreover, we had no information on maternal diet during pregnancy nor did we measure maternal blood pressure. Diet may be a confounding factor since maternal prenatal low energy intake is associated with increased carotid intima-media thickness in later childhood^48^, while it should be noted that our cohort comes from a western, industrialized society with generally sufficient nutrition in the population Additionally, depression and cardiovascular disease may share a common heritable background^49^, which was not addressed by our study.

We have identified maternal prenatal stress as a potential contributor to child cardiovascular health, and added evidence on the importance of early-life factors in directing the cardiovascular health trajectories with plausible reflections into adulthood^50^. Importantly, prenatal stress is a modifiable factor since psychological interventions directed to pregnant mothers and their spouses are potentially effective at alleviating prenatal psychological distress symptoms ^51^. Our findings add important knowledge to the literature of potential cardiovascular disease risk factors in the general population, and especially in males among whom these conditions are prevalent public health issues^52^. Prenatal depressive symptoms as well as pregnancy-anxiety are common during pregnancy. Thus, our results point out the need for intervention studies focusing on early identification and treatment of maternal psychological distress. Designing such studies would show light on the possibilities to improve offspring health through maternal preventive interventions which, according to the developmental origins of health and disease theory, would be most impactful when targeted in early life^50^.

## Conclusions

Our results suggest that pregnancy-specific anxiety is associated with thicker aortic intima-media in 2-3-month-old infants. On the other hand, constantly elevated, mild to moderate depressive symptoms are associated with thicker aortic intima-media in a sex-specific manner, i.e. depressive symptoms are related to thicker aortic intima-media in infant boys only. Overall, this would indicate that maternal prenatal psychological distress, a form of prenatal stress, is related to cardiovascular structure in infancy. Importantly, prenatal psychological distress is a potentially modifiable risk factor and, thus, an interesting intervention target in efforts to improve cardiovascular health and influence disease risks in the population.

## Data Availability

Due to Finnish national legislation, the metadata cannot be made available online, but data can potentially be shared with Data Transfer Agreement as part of research collaboration. Requests for collaboration can be sent to the Board of the FinnBrain Birth Cohort Study; please contact Linnea Karlsson.

## Acknowledgements

We thank FinnBrain Birth Cohort Study families for participating and FinnBrain Birth Cohort Study and Research Centre of Applied and Preventive Cardiovascular Medicine personnel for their efforts.

## Ethical Standards

The authors assert that all procedures contributing to this work comply with the ethical standards of the relevant national guidelines on human experimentation (please name) and with the Helsinki Declaration of 1975, as revised in 2008, and has been approved by the institutional committees of the Hospital District of Southwest Finland

## Appendices

**Supplementary Table 1.**
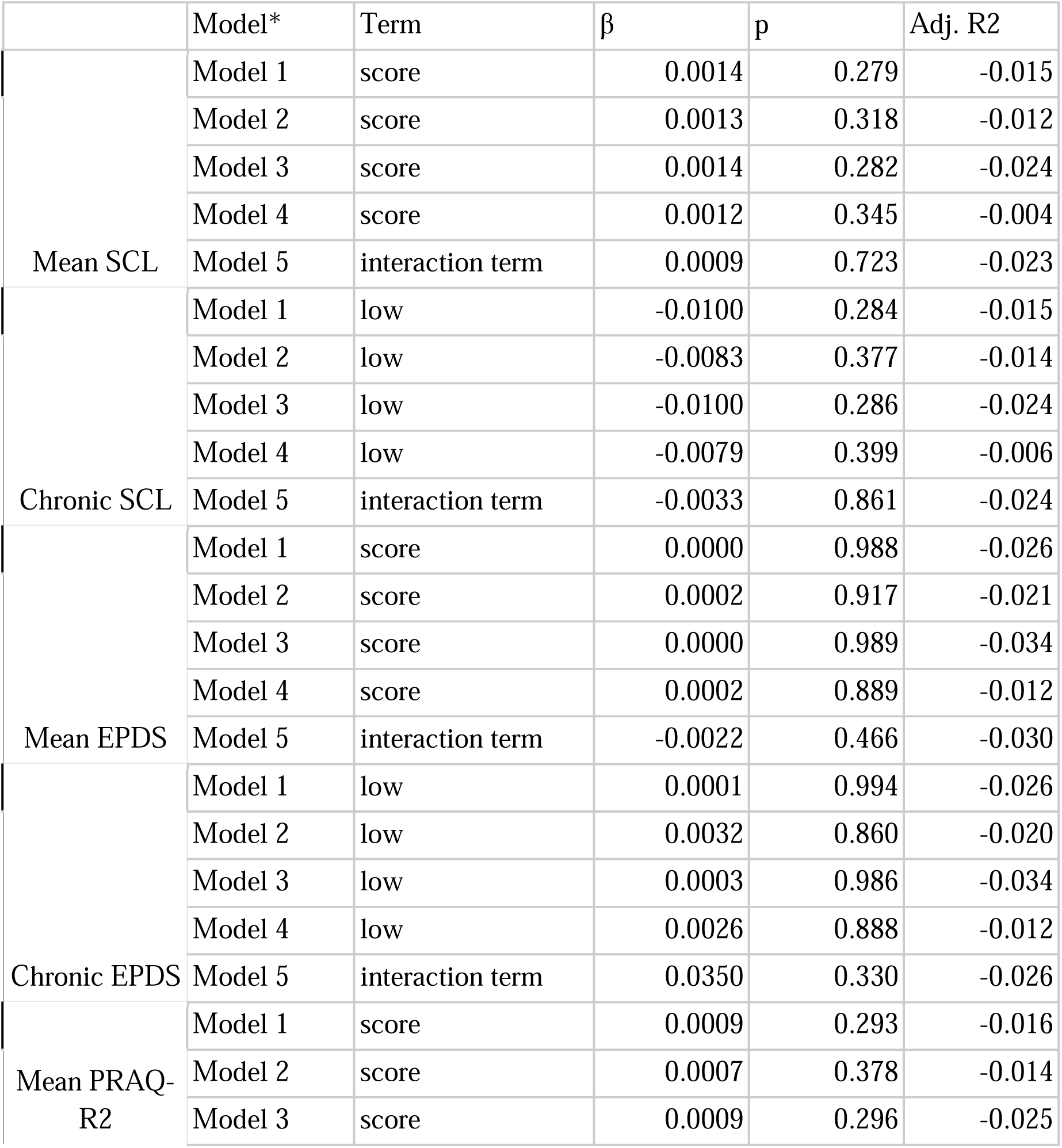
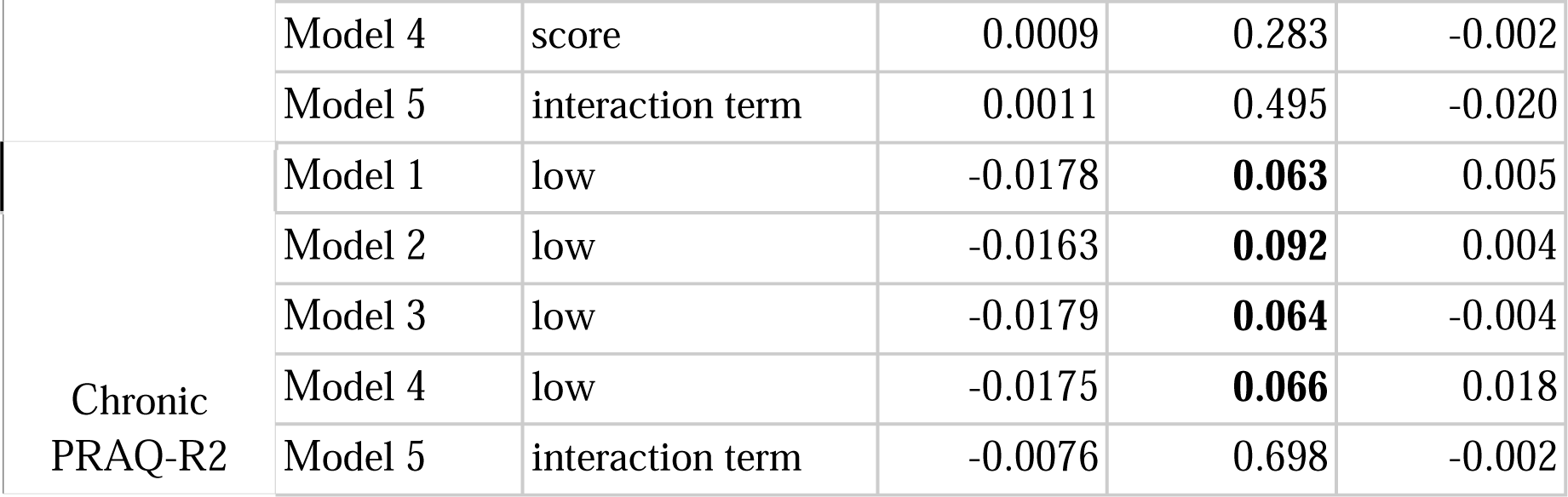
Sensitivity analysis with sample limited to subjects with all symptom and covariate data available (n=124).

